# Assessing food acquisition, preparation, and consumption practices in South Asia: A systematic review of assessment tools

**DOI:** 10.1101/2025.01.06.25320042

**Authors:** Sharvari Patwardhan, Morgan Boncyk, Rasmi Avula, Christine Blake, Fahmida Akter, Jai K Das, Renuka Silva, Purnima Menon, Samuel Scott

## Abstract

Assessing behaviors related to food choice at individual- and household-levels is essential for improving household diets, but assessment tools are limited. We conducted a systematic review to identify gaps in existing assessment tools for food acquisition, preparation, and household consumption practices in South Asia, wherein diets are rapidly changing, and triple burden of malnutrition is emerging.

Systematic search of three academic databases (PubMed, Scopus, and Web of Science Core Collection) using pre-defined keywords were undertaken to identify studies assessing food acquisition, food preparation, and household consumption practices in South Asia, published in English between 2000 and December 2023. Following PRISMA guidelines, two reviewers independently screened titles, abstracts, and full texts based on the inclusion criteria, and extracted data on study characteristics and the assessment tools used to examine the food choice behaviors.

Of 11,288 unique articles identified, 46 were included for synthesis. Food acquisition behaviors were assessed by 25 studies, food preparation by eight studies and household consumption practices by 26 studies. Most studies used quantitative methods (n=30), some used qualitative (n=13), and few used mixed methods (n=3), and varied by type of behavior assessed. Likert scales were the most widely used tools of quantitative assessments, while semi-structured interviews were the most common for qualitative assessments. Across the 46 studies, 59 different tools were used to assess food-related behaviors and only 14 studies claimed using validated tools and many studies did not include the full tool in the text or supplement (n=22).

Our review highlights the need for expanding food choice behavior assessments to include the less-studied populations such as exploring young children and adolescents’ food choice behaviors and developing a contextually adaptable repository of validated tools to advance our understanding of food choice behaviors in various settings.

**Registry**: Open Science Framework Registries

**Registration DOI**: https://doi.org/10.17605/OSF.IO/5GPEF

## INTRODUCTION

Healthy diets play an important role in preventing all forms of malnutrition and diet-related non-communicable diseases (NCDs) ^1^. Individual food choices that shape dietary patterns are important for achieving healthy diets in South Asia, where dietary choices are challenged by poverty, high food prices relative to income, and availability and access constraints^1,2^. Solutions to these diet-related challenges require an understanding of the external food environment along with an examination of food choice behaviors at the household and individual levels^4–6^. Limited knowledge of food choice behaviors and their linkages stems partly from dearth of valid and reliable assessment tools for these behaviors^3^.

Food choice encompasses the processes by which individuals decide what, how, and why to acquire, prepare, allocate, store, consume, and dispose food^7^. This process involves a series of food-related decisions and behaviors that lead to the consumption of particular foods within the context of a specific food environment. Food choice is not limited to food consumption alone. Food choice behaviors can include food acquisition, preparation, allocation, food safety and storage, and waste and disposal behaviors. Food choice is deeply intertwined with expressions of identity, preferences, and socio-cultural values that ultimately shape dietary intake and health outcomes. Such “drivers” of food choice span individual and household levels and are shaped by broader community and macro factors^7^.

Food acquisition, preparation and consumption span the point at which the individual interacts with their food environment to the point when food is consumed. Food acquisition refers to what people acquire, how they acquire it, and where they acquire it, while food preparation refers to actions performed to transform food from raw or partially or fully processed ingredients to a consumable form in the household^7^. Household consumption practices are described according to their patterning (e.g., regularity, skipping, timing), format (e.g., sequence of consumption of food groups), and context (e.g., family meals, engagement in co-occurring behaviors such as watching television)^7^.

Recent methodological advancements to assess food environments in low- and middle-income countries (LMICs) emphasize the external food environment (availability, prices, vendors and product properties, marketing, and regulation) ^4–6^. Assessment of individual and household food choice behaviors within the personal food environment , however, has received less attention^3^. Knowing which behaviors to assess and how, can help guide efforts to improve diets. Providing clarity on which food choice behaviors to measure and the most appropriate tools to measure these behaviors will reduce the need for time-consuming re- creation of data collection protocols.

This systematic evidence and methods review aims to understand whether and how these three food choice behaviors – food acquisition, food preparation, and household consumption practices – have been assessed in South Asia. A secondary aim was to understand the types of food choice drivers being studied (but not how these drivers were assessed). The overarching aim of this work was to identify common themes and best practices for future measurement of food choice behaviors, which is needed to inform interventions to improve diets and wellbeing.

## METHODS

### Data search, screening, and inclusion criteria

This review was registered in OSF (Open Science Framework) Registries (https://doi.org/10.17605/OSF.IO/5GPEF) and followed Preferred Reporting Items for Systematic Reviews and Meta-Analyses (PRISMA) guidelines. Referring to the drivers of food choice and food choice behaviors constructs^7^, an iterative scoping exercise was conducted to identify, test, and refine the appropriate search strings for the three food choice behaviors of interest – food acquisition, preparation, and household consumption practices (**Table S1**). The emphasis was on research that assessed how people eat rather than what people eat. We did not include intra-household food allocation in the review as there is an existing systematic review on this behavior^8^. Although food storage and disposal are important behaviors, we did not include them in the study.

Two authors (SP, MB) ran and filtered the searches in December 2023 in PubMed, Scopus, and the Web of Science Core Collection, which focused on publications in English since the year 2000. The keyword search in the three databases was restricted to the title and abstract. A separate search was conducted for each of the three food choice behaviors. The results from all the searches – for all three behaviors and from all three databases – were pooled and duplicates were removed prior to screening. Search results were imported into Rayyan, a web tool designed for systematic literature reviews^9^, to aid in identifying duplicates and carry out the screening process.

Titles, abstracts, and full texts were screened independently by two reviewers (SP, MB) and disagreements were resolved by a third reviewer (SS, RA, or CEB). All reviewers met weekly during the screening process to ensure consensus and discussed any uncertainties until resolution was achieved.

Articles were included if they were based in a country in South Asia (Afghanistan, Bangladesh, Bhutan, India, Maldives, Nepal, Pakistan, and Sri Lanka), assessed food acquisition, food preparation, or household consumption practices and had at least one driver of food choice at the individual or household level, described the tool used to assess the food choice behavior(s), were peer-reviewed, published in English language from January 2000 to December 2023, and involved a healthy population aged five years or older.

### Data extraction and analysis

Two authors (SP, MB) independently extracted data, and a third reviewer (SS, RA, or CEB) independently reviewed the extracted data to minimize potential bias and verify the accuracy of the extracted data. Data were extracted on authors, title, study country, residence type (rural/urban/peri-urban), study objectives, study population (age and gender), method of assessment, study design, behaviors and drivers assessed, tools used to assess the behavior(s), availability of assessment tool and whether the assessment tool was validated.

Data were grouped according to the classifications of food choice behaviors^7^. Food choice drivers were grouped according to the domains described in Boncyk et al. 2023^7^. Quantitative assessment tools were further categorized as self-administered or interviewer-administered questionnaires and qualitative tools were further categorized as semi-structured interviews, focus group discussions, pile sorting, or photovoice. The availability of questionnaires or tools used to assess the behavior(s) were categorized as ‘descriptions of questions specified’, ‘few questions specified’, or ‘complete tool included’. The article text and supplemental material were reviewed to locate specific questions. Tools were described as validated if the authors directly stated that the tool has been validated within the study context. Validity was not applicable for qualitative tools such as focus group discussions (FGDs).

## RESULTS

### Summary of evidence across food choice behaviors

A total of 22,339 articles were identified. After removing duplicates, 11,288 titles, 562 abstracts, and 167 full texts were screened (**Figure 1**). A total of 46 studies met our inclusion criteria and were included for synthesis. Details on the study population, data collection method, food choice behaviors assessed, and domains of food choice drivers assessed in each of the 46 studies are shown in **Table S2**.

**Figure 1:**
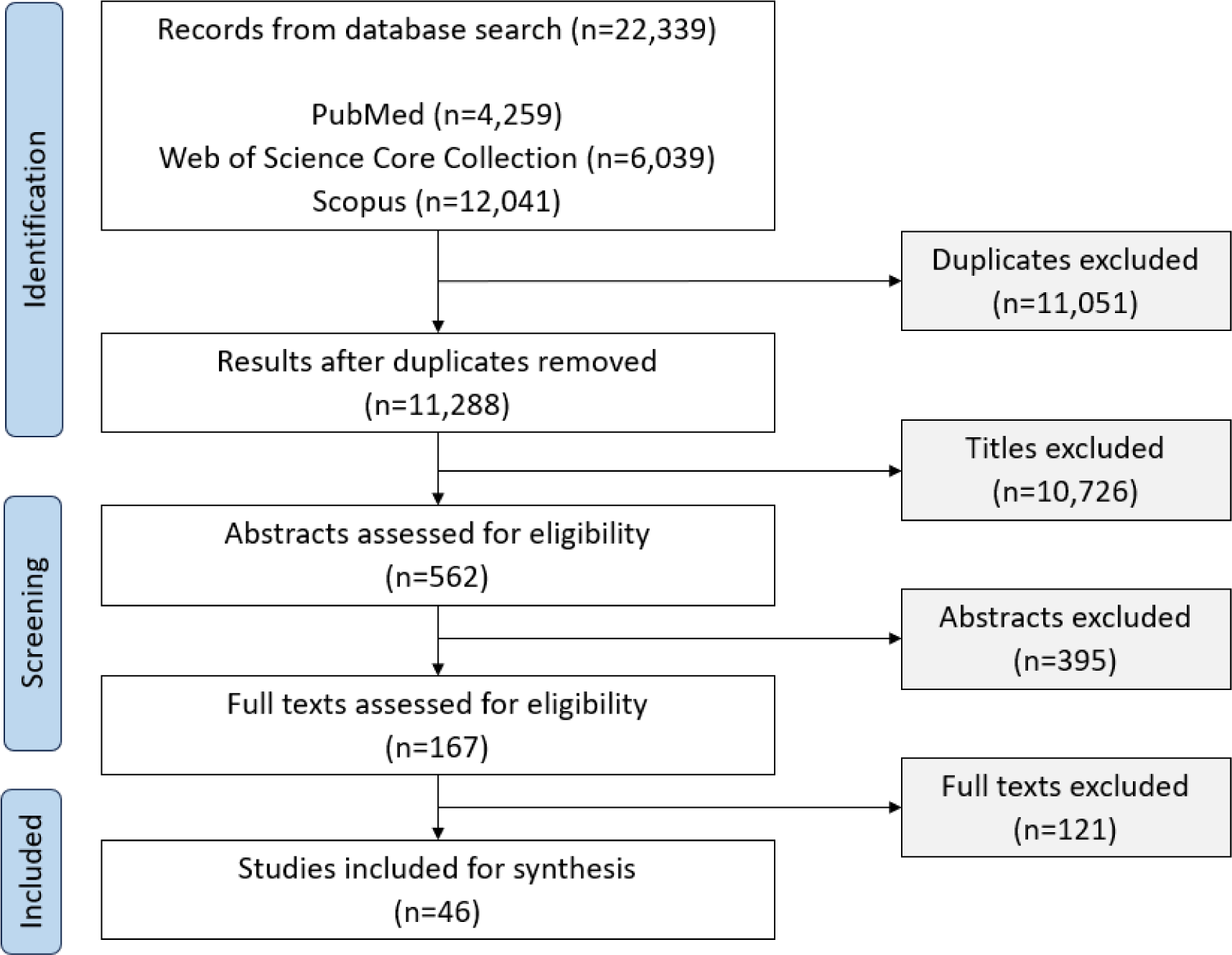
PRISMA-driven flowchart for selection of studies included in the review.

Food acquisition behaviors were assessed by 25 studies, food preparation by eight studies and household consumption practices by 26 studies (**Table 1**). Most studies were from India (n=36), with few from Pakistan (n=4), Bangladesh (n=3), Nepal (n=2), and Sri Lanka (n=1). No studies were identified from Afghanistan, Bhutan, or Maldives and no studies examined multiple countries. Over two-thirds of the studies occurred in an urban setting (n=25), three studies were from a rural setting, one study was from a peri-urban setting and six studies did not specify the region. Most studies included only adults (n=29) while few included only adolescents (n=6), younger children (n=1), or mixed age groups (n=10). Fewer studies examined only females (n=3) or males (n=2) as most examined a mixed-gender population (n=38).

**Table 1:** Summary of the number of included studies by food choice behavior.

|  | Food acquisition | Food preparation | Household consumption practices | No. of unique studies |
| --- | --- | --- | --- | --- |
| **Location** |  |  |  |  |
| *Country* |  |  |  |  |
| Afghanistan | 0 | 0 | 0 | 0 |
| Bangladesh | 1 | 1 | 2 | 3 |
| Bhutan | 0 | 0 | 0 | 0 |
| India | 19 | 6 | 23 | 36 |
| Maldives | 0 | 0 | 0 | 0 |
| Nepal | 1 | 0 | 1 | 2 |
| Pakistan | 4 | 0 | 0 | 4 |
| Sri Lanka | 0 | 1 | 0 | 1 |
| *Residence type* |  |  |  |  |
| Urban | 19 | 5 | 21 | 25 |
| Rural | 6 | 1 | 9 | 3 |
| Peri-urban | 1 | 1 | 1 | 1 |
| Not specified | 4 | 2 | 2 | 6 |
| **Study population** |  |  |  |  |
| *Age* |  |  |  |  |
| Younger children (5-10 years) | 0 | 0 | 2 | 1 |
| Older children/adolescents (10-19 years) | 5 | 2 | 11 | 6 |
| Adults (>19 years) | 21 | 8 | 20 | 29 |
| *Gender* |  |  |  |  |
| Male | 0 | 1 | 1 | 2 |
| Female | 1 | 2 | 3 | 3 |
| Both | 21 | 5 | 22 | 38 |
| **Methods** |  |  |  |  |
| *Quantitative* |  |  |  |  |
| Self-administered questionnaire | 9 | 2 | 7 | 13 |
| Interviewer-administered questionnaire | 11 | 0 | 10 | 17 |
| *Qualitative* |  |  |  |  |
| Semi-structured interviews | 7 | 6 | 7 | 6 |
| Focus group discussions | 2 | 2 | 5 | 2 |
| Pile sorting | 0 | 1 | 2 | 0 |
| Photovoice | 1 | 1 | 1 | 0 |
| *Tool availability* |  |  |  |  |
| Description of questions specified | 15 | 2 | 16 | 33 |
| Few questions specified | 1 | 1 | 2 | 4 |
| Complete tool specified | 9 | 5 | 8 | 22 |
| *Tool validity* |  |  |  |  |
| Validated | 5 | 1 | 8 | 14 |
| Not validated | 14 | 1 | 8 | 23 |
| Not applicable (e.g., qualitative FGDs) | 6 | 6 | 10 | 22 |
| Column total | 25 | 8 | 26 | 46 |

Note: Categories are not mutually exclusive.

| Legend |  |  |  |  |  |
| --- | --- | --- | --- | --- | --- |
| No of studies | 0 | 1-5 | 5-9 | 10-14 | ≥15 |

Thirty studies used quantitative methods, 13 studies used qualitative methods, and 3 studies used mixed methods for their analyses. Quantitative tools included questionnaires (self- and interviewer-administered) with various scales and qualitative tools included semi-structured interviews and FGDs. Across the 46 studies, 59 different tools were used, only 14 of which were reported by the authors to be validated. The complete tool was provided in 22 studies, with the remaining studies only describing the questions or providing a few questions.

### Tools used to assess food choice behaviors

#### Assessment of food acquisition behaviors

Twenty-five studies described food acquisition behaviors in terms of frequency of purchases (e.g., frequency of buying outside food, buying from the school canteen)^10–12^, purchase of specialty foods (e.g., supersized foods, halal food, organic food)^13–26^, influence of the COVID-19 pandemic on food buying behavior (e.g., changes in food shopping behavior, market frequency)^27–30^, intra-household acquisition practices (e.g., where and who acquires food)^31–32^, and online purchasing (e.g., pattern of online food orders, consumption occasions for ordering online food)^33–34^ (**Table 2**). Frequency of food purchases were assessed using both quantitative (questionnaires)^10,12^ and qualitative (FGDs)^11^ approaches. Most studies assessing purchase behavior for specialty foods (in terms of purchase frequency, purchase patterns, purchase preferences) used quantitative tools; Likert scales^13–15,17–18,22,24^ were most commonly used by interviewer-administered surveys^16,19,25^ and online questionnaires^23^. Few studies used qualitative tools such as semi-structured interviews^21,26^ including open-ended questions^20^ to assess purchase behavior. Changes in food acquisition behaviors during the COVID-19 pandemic were assessed primarily using quantitative self-administered questionnaires^27–29^ while only one study conducted qualitative FGDs^30^. Studies examined intra-household food acquisition practices using qualitative interviews^31–33^ which were supplemented with photovoice^31^ and quantitative surveys^32^ administered via phone and mail^34^. Likert scales were the most commonly used quantitative tools and semi-structured interviews were the most commonly used qualitative tool.

**Table 2:** Summary of tools used to assess food acquisition behaviors.

| Assessment tool | Population and context | Description of behaviors assessed | Description of scale/questions |
| --- | --- | --- | --- |
| **Frequency of food purchases** |  |  |  |
| Qualitative: Focus Group Discussions (FGDs) | Adolescents and their caregivers living in urban slums (urban, India) | Frequency of buying outside food | How frequently do your family members give you money to buy outside food? What foods do you buy from the money given by the family? (Chopra et al. 2021) |
| Quantitative: Survey (Interviewer-administered questionnaire) | Low- and middle-income households (urban and rural, India) | Purchasing behaviors | The questionnaire collected information about consumers' purchase and physical access to various food products and perceptions on the promotional aspect of food (Ynion et al. 2021). |
| Quantitative: Survey (Self-administered questionnaire) | School-going adolescents (urban, India) | Frequency of buying from the school canteen | Do you eat at the school canteen or buy foods/beverages at school? If yes, then how often...? Why do you buy these foods? Response options included taste, price, availability, and convenience (Moitra, P. & Madan, J. 2022). |
| **Purchasing behavior of specific foods** |  |  |  |
| Qualitative: Interviews (Semi-structured interviews) | Consumers of supersized foods (urban, Pakistan) | Supersized foods purchase behavior | When was the last time they ordered supersized food/drink? What causes consumers to purchase supersized food? Does larger package or supersized food within an assortment of product sizes reflect status? (Qazi et al. 2022) |
|  | Consumers of western imported foods (urban, Pakistan) | Western imported foods purchase behavior | Specific questions not mentioned (Bukhari et al. 2022). |
|  | Consumers of western imported foods (urban, Pakistan) | Buying behavior towards edible oil and vanaspati ghee | Preference of respondent takes value 1 if oil is preferred and vanaspati ghee otherwise. Preference of oil assessed against health concern, or due to taste / aroma (1 if Yes and No otherwise). (Ullah et al. 2020) |
| Quantitative: Survey (Interviewer-administered questionnaire) | Adults (urban, India) | Frequency of purchasing probiotics | Questions helped to assess acceptance, brand awareness and willingness to purchase probiotic products (Arora et al. 2021). |
|  | Adolescents (urban, India) | Fast food buying behavior | Factors influencing buying behavior towards fast food were recorded on a three-point Likert scale (3 =most influencing factor, 2= moderately influencing factor, and 1 = least influencing factor). (Malushte et al. 2022) |
|  | Consumers (urban, Bangladesh) | Halal food purchase behavior | How many times would you say to purchase halal food per month? How long have you been buying halal foods? (Ashraf, M. A. 2019) |
|  | Consumers (urban, India) | Meat purchase patterns | Information collected on mode of purchase (from shop, slaughter in home or front of eyes, frozen/supermarket) and preferred product type (Kiran et al. 2018). |
|  | Consumers (urban, India) | Purchase Behavior for Health and Wellness Food Products | How often do you purchase organically grown produce or other organic food products? (Budhathoki, M., & Pandey, S. 2021) |
|  | Consumers of organic food (urban, Nepal) | Purchase frequency of organic foods | Factors affecting buying behavior were recorded on a five-point Likert scale (1 = not at all important and 5 = extremely important) (Ali et al. 2021). |
|  | Experts in the food industry (India) | Purchase process of organic foods | The questionnaire involved a mix of structured and half-structured questions. Respondents evaluated the importance of 19 variables in the consumers' purchase process on a 1-5 scale (Chakrabarti, S. 2010) |
| Quantitative: Survey (Self-administered questionnaire) | Consumers representative of households (urban and rural, India) | Non-packaged non-branded rice purchase behavior | Questions about frequency of purchase, quantity purchased, source of purchase, and mode of purchase (Prasad R. & Umesh S. 2016). |
|  | Young consumers/ students (urban and rural, India) | Purchase frequency of organic foods | How often have you shopped for organic foods in the last three months (1 = "never"; 5 = "at every opportunity")? How many organic foods have you bought over the last three months (1= "none at all"; 5 = "a great deal")? (Matharu et al. 2023) |
|  | Adult respondents interested in organic foods (urban, India) | Purchase frequency of organic foods | Responses to the statement "Occasionally, I purchase organic foods" were recorded on a five-point Likert scale, from agree to disagree (David et al. 2020). |
|  | Young consumers/ students (urban and rural, India) | Purchase frequency of organic foods | All constructs about factors influencing organic food purchase were measured on a five-point Likert scale, ranging from 1 for "Strongly disagree" to 5 for "Strongly agree" (Matharu et al. 2022). |
| **Changes in food acquisition behavior due to the COVID-19 pandemic** |  |  |  |
| Qualitative: Interviews (Semi-structured interviews) | Female food gatekeepers/ those responsible for feeding their family members, COVID19 pandemic (urban, India) | Food shopping behavior | Can you explain about your food shopping experience before and during lockdown? From which sources do you usually procure food on a regular basis (before and during lockdown)? (Menon et al. 2022) |
| Quantitative: Survey (Self-administered questionnaire) | Adults, COVID-19 lockdown (India) | Market frequency | Information collected on the frequency of going to market during the lock down period (Samanta et al. 2022). |
|  | Adults, COVID-19 pandemic (urban and rural, India) | Food buying behavior | How much COVID-19 reduced buying frequency for food purchase? Has COVID-19 changed buying behavior towards online purchases (1 if people changed buying behavior, and 0 otherwise)? (Shahzad et al. 2022) |
|  | Consumers, COVID-19 lockdown (India) | Meat buying behavior | Questionnaire collected information on meat procurement source, type of meat/meat product purchased, meat available even during the lockdown period, and the quantity of meat procured during COVID19 lockdown to the normal situation (Rahman et al. 2021). |
| **Intra-household food acquisition practices** |  |  |  |
| Qualitative: Interviews (Semi-structured interviews) | Peri-urban households (peri-urban, India) | Food acquisition practices | In-depth interviews about current food acquisition practices, and intra- household food acquisition (Turner et al. 2022). |
|  | Low- and middle-income households (urban, India) | Where and who acquires food | Key informants (food vendors) were interviewed to explore food purchasing behavior of the households (Pradhan et al. 2013). |
| Qualitative: Photovoice (Participant photographs) | Peri-urban households (peri-urban, India) | Food acquisition practices | Participants photographed their food acquisition practices over a three-day period. With these, photo-elicitation follow-up interviews were conducted with participants (Turner et al. 2022). |
| Quantitative: Survey (Interviewer-administered questionnaire) | Low- and middle-income households (urban, India) | Where and who acquires food | The structured survey schedule focused on a) usual food acquisition in the household, b) household decision-making on food purchases, c) the usual food purchaser, d) location of where food purchases were made (Pradhan et al. 2013). |
| **Online food purchases** |  |  |  |
| Qualitative: Interviews (Semi-structured interviews) | Adults, COVID19 lockdown (India) | Pattern of online food orders | A subset of participants who were found to be food addicts based on the Yale Food Addiction Scale were asked - During this lockdown phase, have you ordered ready-to-eat (cooked) food online? If yes, how many times (Yes/No/Other). (Das et al. 2021) |
| Quantitative: Survey (Self-administered questionnaire) | Adults, COVID19 lockdown (India) | Pattern of online food orders | Yale Food Addiction Scale used to assess whether respondents were food addicts. The questionnaire asked if participant's eating behavior caused significant distress and also whether they experienced significant problems in being able to function effectively due to their food or eating habits. Responses recorded on a five-point Likert scale (Das et al. 2021). |
|  | Online food delivery customers (urban, India) | Recurring consumption occasions for ordering online food and factors impacting the same | The questionnaire had sub-sections viz. demographic profile, consumption and spending pattern, factors impacting online food ordering, and overall satisfaction (Vinish et al. 2021). |
| Quantitative: Survey (Interviewer-administered questionnaire) | Online food delivery customers (urban, India) | Recurring consumption occasions for ordering online food and factors impacting the same | The questionnaire had sub-sections viz. demographic profile, consumption and spending pattern, factors impacting online food ordering, and overall satisfaction (Vinish et al. 2021). |

#### Assessment of food preparation behaviors

Eight studies^11,27,30–31,33,35–37^ described assessment of food preparation behaviors, including intra-household distribution of food preparation responsibilities^31,35^ and cooking habits (e.g., the type of food typically prepared, time spent in the kitchen)^11,27,30,36–37^ (**Table 3**). Intra-household distribution of food preparation responsibilities was assessed using qualitative tools such as semi-structured interviews^31,35^ and photovoice^31^. Although two studies conducted semi-structured interviews, their approach was different - one study focused on general intra-household preparation^31^ while the other study enquired about how food preparation and cooking responsibilities differ between household members^35^. Cooking habits were examined using qualitative interviews^30,33,37^, FGDs^11,37^, pile sorting^36^, and quantitative surveys^27^. Most studies assessing food preparation behaviors used qualitative tools such as semi-structured interviews. Quantitative self-administered questionnaires were implemented only by two studies^27,33^.

**Table 3:**
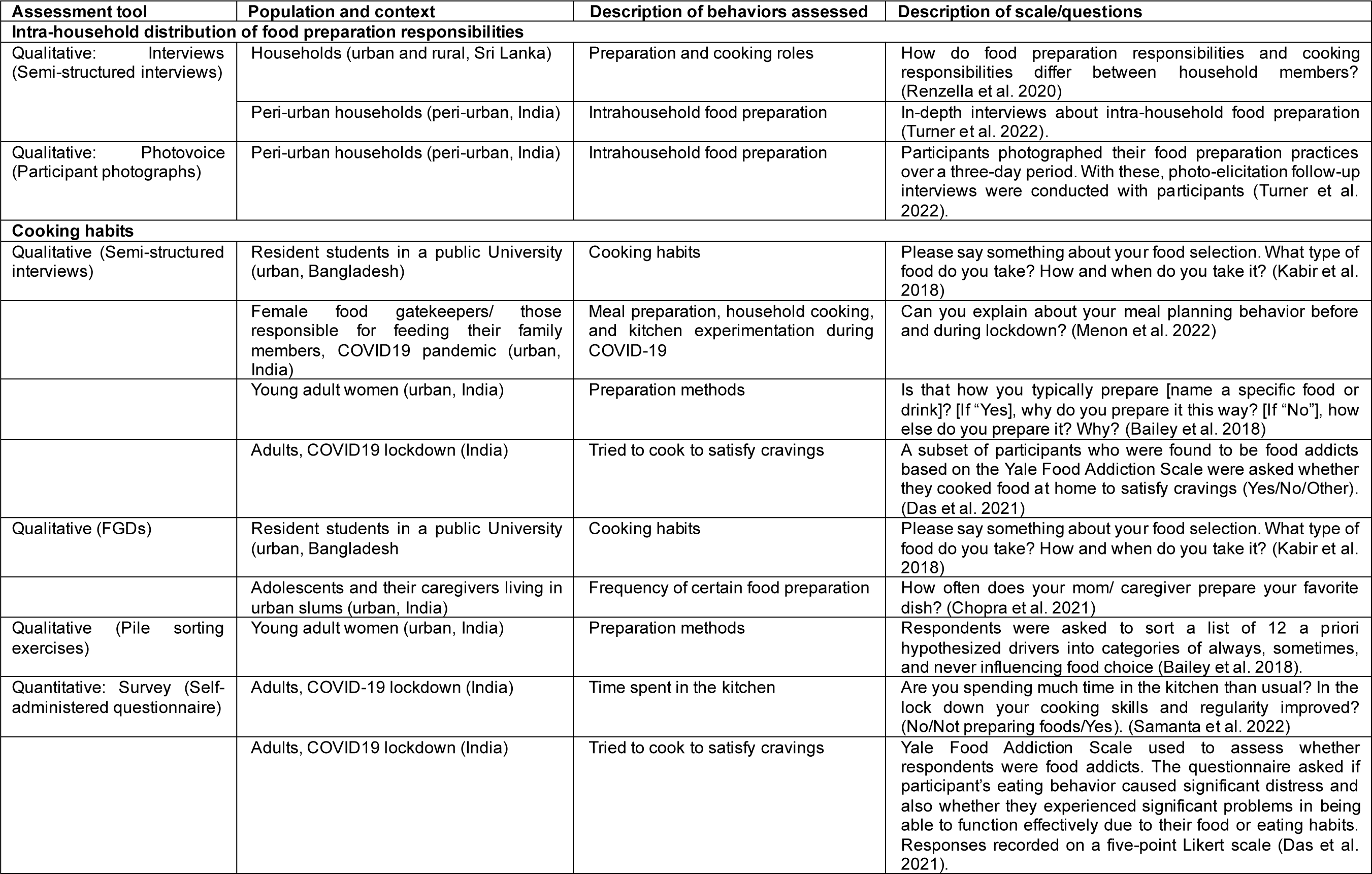
Summary of tools used to assess food preparation behaviors.

#### Assessment of household consumption practices

Twenty-six studies described household consumption practices, including intra-household food distribution and consumption^30,32,38–41^, foods consumed during various eating occasions^12,36^, eating habits^31,42,44–46^, eating behaviours^37,43^, snack consumption patterns^10,11,47–51^, and eating out behavior^52–55^ (**Table 4**).

**Table 4:** Summary of tools used to assess household consumption practices.

| Assessment tool | Population and context | Description of behaviors assessed | Description of scale/questions |
| --- | --- | --- | --- |
| **Intra-household food allocation practices** |  |  |  |
| Qualitative (Semi-structured interviews) | Low- and middle-income households (urban, India) | Food distribution patterns | In-depth interviews about intra-household food distribution patterns (Pradhan et al. 2013). |
|  | Adolescent females from low-income households (urban and rural, Bangladesh) | Food allocation | Specific interview guidelines not mentioned (Blum et al. 2019). |
|  | Female food gatekeepers/ those responsible for feeding their family members), COVID19 pandemic (urban, India) | Family mealtime | Can you tell me about your social eating dynamics (i.e. family mealtime) before and during lockdown? (Menon et al. 2022) |
| Qualitative (FGDs) | Families in the community (rural, India) | Eating order | Who eats first, and who eats last? What happens when the food is finished and the woman of the household hasn't yet eaten? Does she go back and cook more food? How else might she get food? (Neogy, S. 2010) |
| Qualitative (Pile sorting exercises) | Families in the community (rural, India) | Eating order | At the group and household level, flash cards were used to guide the facilitator in general discussion points (Neogy, S. 2010). |
| Qualitative (Free listing exercises) | Adolescent females from low-income households (urban and rural, Bangladesh) | Food allocation | Free listing exercises conducted to identify foods eaten during meals and eating patterns (Blum et al. 2019). |
| Quantitative: Survey (Interviewer-administered questionnaire) | Children and their parents from educationally backward areas (rural, India) | Whether girls eat after boys in the household | Parents asked to what extent they agreed with statements reflecting gender roles and attitudes. Responses collected on a three-point Likert scale (Agree/ Agree to an extent/ Disagree) (Ghatak et al. 2024). |
|  | Low- and middle-income households (urban, India) | Food distribution patterns | Questions on intra-household food distribution patterns (Pradhan et al. 2013). |
|  | Adults highly involved in decision making in the family (urban and rural, India) | Urban-rural differences in household food consumption | In which form do you usually buy wheat; frequency of consumption of selected convenience food products; media that influences food products. Importance of nine food choice motives scored between 1-7 (Mor, K. & Sethia, S. 2018). |
| **Foods consumed during various eating occasions** |  |  |  |
| Qualitative (Semi-structured interviews) | Young adult women (urban, India) | Foods consumed during different eating occasions | Is this what you typically eat at this time of day? [If "No"], how is it different? (Bailey et al. 2018) |
| Qualitative (FGDs) | Low- and middle-income households (urban and rural, India) | Frequency of eating occasions | Statements collated relating to food quality attributes for each eating occasion (Ynion et al. 2021). |
| Qualitative (Pile sorting exercises) | Young adult women (urban, India) | Foods consumed during different eating occasions | Respondents asked to sort a list of 12 a priori hypothesized drivers as always/ sometimes/ never influencing food choice (Bailey et al. 2018). |
| Quantitative: Survey (Interviewer-administered questionnaire) | Low- and middle-income households (urban and rural, India) | Frequency of eating occasions | Respondents asked to evaluate predefined statements relating to food quality attributes for each eating occasion (i.e., breakfast, morning snacks, lunch, afternoon snacks, and dinner) using a 5-point importance scale. Information regarding dishes consumed during daily eating occasions and the corresponding frequency of consumption of each dish was collected (Ynion et al. 2021). |
| **Eating habits** |  |  |  |
| Quantitative: Survey (Semi-structured interviews) | Peri-urban households (peri-urban, India) | Eating practice | In-depth interviews about intra-household food consumption practices (Turner et al. 2022). |
| Qualitative: Photovoice (Participant photographs) | Peri-urban households (peri-urban, India) | Eating practice | Participants photographed their food environment and consumption practices over a three-day period. With these, photo-elicitation follow-up interviews were conducted with participants (Turner et al. 2022). |
| Quantitative: Survey (Interviewer-administered questionnaire) | Young adults (urban, India) | Irregularity in eating habits | Participants asked whether they regularly consume breakfast/lunch/evening food and the reasons for any irregularity in food habit (Patel et al. 2010). |
|  | Adolescents of upper socio-economic status in public schools (urban, India) | Meal skipping | Questionnaire included sections on demographic data, dietary habits and exercise pattern (Aggarwal et al. 2006). |
| Quantitative: Survey (Self-administered questionnaire) | University students (urban, Nepal) | Eating practice and food choice | Food Choice Questionnaire contained 20 items designed to assess the importance of nine factors. Each item scored on a 4-point importance scale. Eating practice was based on a scale of two ("1 = good eating practice", and "0 = otherwise"). (Dahal et al. 2022) |
| | Adolescent school children (urban, India) | Eating habits | Frequency of major meals/day (1–2/3 />3 times), frequency of snacking ($\leq 3/4$/>4 times), history of skipping meals (never/sometimes/often) and history of eating outside home (never/sometimes/often) were recorded. Overall eating habit determined by scoring the relatively poorer eating habits (based on inappropriate frequency of intake, skipping meals and eating fast foods). (Kumar et al. 2017) |
| **Eating behaviors** |  |  |  |
| Qualitative (Semi-structured interviews) | Resident students in a public University (urban, Bangladesh) | Eating behaviors | What/ how/ why/ where do you eat? Who serves you and how? What affects your eating and how? Please say something about your food selection. What type of food do you take? Please discuss elaborately (when, how, why, and why not?). (Kabir et al. 2018) |
| Qualitative (FGDs) | Resident students in a public University (urban, Bangladesh) | Eating behaviors | How and what do you eat? In your opinion, what aspects/issues/elements affect your eating in and around your university? (Please discuss elaborately when, how, why and why not?). (Kabir et al. 2018) |
| Quantitative: Survey (Self-administered questionnaire) | School-going children (India) | Eating behaviors | 8 dimensions of eating style in children assessed (responsiveness to food, enjoyment of food, satiety responsiveness, slowness in eating, fussiness, emotional overeating, emotional under eating, and desire for drinks). A total of 35 questions asked with 5 options (Never/ Rarely/ Sometimes/ Often/ Always). Each question related to a particular dimension (Semar, R. & Bakshi, N. 2022). |
| **Snack consumption patterns** |  |  |  |
| Qualitative (Semi-structured interviews) | Consumers of various age groups (urban and rural, India) | Frequency, reasons, timings of snack consumption | The questionnaire included 126 questions for 25 food groups. Consumption pattern (including frequency of consumption) of routine meals and snack foods for the past month, their timing, habits related to skipping meals, and factors associated with choice of different snacks were assessed (Roy et al. 2021). |
| Qualitative (FGDs) | Adolescents and their caregivers living in urban slums (urban, India) | Fast food consumption | How frequently do your family members give you money to buy outside food? What foods do you buy from the money given by the family? How often do you eat with your family/ other families/ elsewhere in the community? How do your friends and family affect your choice of food? (Chopra et al. 2021) |
| Quantitative: Survey (Interviewer-administered questionnaire) | Medical students (India) | Regularity of eating fast foods | Questions regarding respondents' food habits and the barriers that prevented them from maintaining healthy dietary habits (Mohanty et al. 2017). |
|  | Adults (urban and rural, India) | Context for snack consumption | Multiple choice questions were designed to understand the preferences for activities done while eating snacks, and top reasons for choosing snacks. Participants selected 3 options of all and ranked them in the order of preference (Ganpule et al. 2023). |
| Quantitative: Survey (Self-administered questionnaire) | Pre-university students (urban and rural, India) | Fast food consumption patterns | Participants answered 10 questions to assess their knowledge and practice of fast food consumption (1 point for the right answer and 0 for a wrong answer). Overall score ranged from 0–10 (Khongrangjem et al. 2018). |
|  | School-going adolescents (urban, India) | Context for snack consumption | In the last 7 days, how many days did you perform the following activities such as watching television while eating? The response options, 'never' to 'always, indicating < 1/week to 6–7 times/ week, were scored on a five-point scale from 0 to 4. Higher scores indicated a higher frequency of indulging in a specific eating habit (Moitra, P. & Madan, J. 2022). |
|  | Adults, COVID19 lockdown (urban, India) | Context for snack consumption | The total number of meals consumed before and during lockdown, whether participants consumed snacks at those meals, and reasons for change in snacking pattern during lockdown were assessed using a pre-structured list of beverages and snacks (to examine intake during and before lockdown) (Bhol et al. 2021). |
| **Eating out behavior** |  |  |  |
| Qualitative (FGDs) | Adults of various socioeconomic groups (urban, India) | Frequency of eating out | What is your opinion about eating out? (Probe questions - Do they have more or less fat, sugar and salt?) What do you think are the reasons for fast-food preference and dining out? How frequently do you eat out? (Kaur et al. 2020) |
| Quantitative: Survey (Interviewer-administered questionnaire) | Children and their parents (urban and rural, India) | Frequency of eating out | The questionnaire enquired about preferences of eating out and if participants were affected by/ changed their buying behavior in light of the information they interpreted from food labels. Another section included perceived concerns about the tv ads for children's food (Verma et al. 2023). |
|  | Consumers (urban, India) | Frequency of eating out | Questionnaire included questions related to the frequency of eating out in a month, preference of cuisine between Indian or Chinese, vegetarian and non-vegetarian, spending per visits and advertisement; recorded on a nominal scale (Khan et al. 2021). |
| Quantitative: Survey (Self-administered questionnaire) | Street food consumers (urban, India) | Eating food sold by street vendors | Word of Mouth scale, Food Neophobia scale, and Intention to Consume scale used. Responses to statements were recorded on a five-point Likert scale (extremely unlikely to extremely likely). (Khanna et al. 2022). |

Qualitative semi-structured interviews^30,32,40^ FGDs^38^, and systematic data collection methods^38,40^ were the most common approaches to assess intra-household food distribution and consumption followed by quantitative Likert scales^39^ and importance scores^41^. A combination of qualitative interviews^36^, pile sorting^36^, FGDs^12^, and quantitative survey tools^12^ were used to examine foods consumed during various eating occasions. For instance, in the pile sorting exercises, respondents were requested to sort a list of a-priori hypothesized drivers into piles of always/sometimes/never influencing food choice^36^. Eating habits and behaviors were assessed primarily using quantitative tools^42–46^; some articles used qualitative interviews^37^, photovoice^31^, and FGDs^37^. A higher proportion of studies assessing snack consumption patterns and eating out behavior used quantitative tools^10,47–48,50–52,54–55^ followed by FGDs^11,53^ and qualitative interviews^49^. Out of the two studies conducting FGDs, one enquired about how frequently adolescents were given money to buy outside food and which foods were bough using that money^11^, while the other study enquired about the frequency of eating out and what the reasons might be for dining out^53^. A combination of quantitative and qualitative tools were used to assess household consumption practices. Quantitative questionnaires followed by semi-structured interviews and FGDs were the most commonly used tools.

## DISCUSSION

This review of studies from South Asia examined the tools used to assess three food choice behaviors and showed that most evidence exists for household consumption practices, followed by food acquisition behaviors and a few studies on food preparation. Most studies were conducted in India and two-thirds of the studies assessed these food related behaviors for adults only. Most of the included studies used quantitative methods. Likert scales were the most widely used quantitative tools, while semi-structured interviews were the most common qualitative tools. There was large heterogeneity between studies in terms of methods and tools used to assess food choice behaviors, limiting comparison of findings across studies.

The aforementioned gap in existing literature has implications in planning culturally sensitive, contextually appropriate interventions and developing policies aimed at improving dietary practices and nutritional outcomes. The lack of standardized tools to measure food choice behaviors among children and in rural areas complicates efforts to accurately assess and compare food preparation practices across diverse settings. Therefore, further research is needed to be conducted in diverse rural settings and population subgroups to understand context-specific roles and challenges, and specially to explore children’s involvement in food preparation which may establish long-term favorable dietary habits among them.

Only a few studies combined quantitative surveys with qualitative interviews or FGDs to triangulate findings and gain a comprehensive understanding of food choice behaviors. Even when assessing similar food choice behaviors using similar methods, the specific tools used differed. For instance, when assessing purchase frequency of organic foods using 5-point Likert scales, one study asked about how often respondents shopped for organic foods while the other study asked respondents level of agreement to a statement about occasionally purchasing organic foods.

Moreover, there was a lack of studies using validated tools. Developing and implementing new tools to assess food choice behaviors that have been previously examined in a different setting is a time and resource-consuming exercise. Hence, efforts are needed to develop a repository of validated tools that can be adapted to different contexts to optimize research resources and maximize comparability between different studies. Gathering data using valid and effective tools provides policymakers with evidence-based guidance for supporting individuals and communities during challenging times.

The key strength of this study is that it provides a comprehensive review of tools used to assess three crucial food choice behaviors - food acquisition, food preparation, and household consumption practices - in South Asia. However, several included studies considered specific foods, such as non-packaged non-branded rice, unhealthy snacks, probiotic food and beverages, halal food, rather than general food choices, which may limit generalizability. Additionally, some studies examined food purchasing behaviors during the COVID-19 pandemic, which may not be reflective of usual practices. Furthermore, the review may have missed relevant articles due to limitations in the type of publication (e.g., exclusion of gray literature), or language restrictions (non-English articles were excluded). Lastly, the lack of consistency and comparability in measurement tools across studies posed challenges in drawing conclusions about the tools used for different types of food choice behaviors.

Food choice behaviors were assessed using various tools, often tailored to specific behaviors, which are valuable for capturing their complexity. While this diversity enhances understanding, inconsistent approaches to measuring the same elements across studies create significant heterogeneity, limiting opportunities for comparability and cumulative understanding. With multiple tools to measure the same element, many tools lack thorough development, validation, and accessibility, further complicating high-quality measurement. Over half the included studies did not report the complete set of tools used, hindering researchers from reproducing, adapting, and improving existing methods. Efforts have started to develop a centralized repository of existing measures, instruments, and protocols to assess food choice behaviors but remains incomplete and requires further refinement to maximize its utility^1^. There is a need for greater standardization across contexts and clear documentation of methods and instruments used. Journals should require the inclusion of assessment tools for newly developed measures to ensure rigor and accessibility. Adapting and validating existing tools, rather than creating new ones, can improve efficiency, continuity, and comparability, enabling researchers to focus on advancing our understanding of food choice behaviors.

## Data Availability

All data produced in the present study are available upon reasonable request to the authors

## SUPPLEMENTARY TABLES AND FIGURES

**Table S1:** Search strings used after initial scoping.

|  | Keyword |
| --- | --- |
| Food acquisition | ("food purchas*" OR "food buy*" OR "food shop*" OR "food acqui*" OR "food vendor*" OR "food"<br>AND<br>("famil*" OR "home" OR "household*" OR "individual*")<br>AND<br>("barter*" OR "exchang*" OR "trad*" OR "grow*" OR "produc*"))<br>AND<br>("Afghanistan" OR "Bangladesh" OR "Bhutan" OR "India" OR "Maldives" OR "Nepal" OR "Sri Lanka" OR "Pakistan") |
| Food preparation | ("cook*" OR "food prepar*" OR "meal prepar*" OR "meal plan" OR "meal plans" OR "meal planning" OR "home prepar*" OR "homemade" OR ("food" OR "meal*" OR "ingredient" OR "cook*")<br>AND<br>("bake" OR "baking" OR "fry" OR "frying" OR "clean*" OR "chop*" OR "boil*" OR "broil*" OR "grill*"))<br>AND<br>("Afghanistan" OR "Bangladesh" OR "Bhutan" OR "India" OR "Maldives" OR "Nepal" OR "Sri Lanka" OR "Pakistan") |
| Household eating patterns | ("eating episode" OR "food consumption behavior" OR ("meal*" OR "breakfast" OR "lunch" OR "dinner" OR "dessert" OR "snack*" OR ("consum*" AND "food") OR "meal*" OR "eat" OR "eating")<br>AND<br>("pattern*" OR "sequence" OR "speed" OR "location" OR "skip*" OR "on-the-go" OR "take out" OR "away from home" OR "home*" OR "travel*" OR "share" OR "shares" OR "sharing" OR "shared" OR "separate" OR "sit" OR "sitting" OR "fami*" OR "time" OR "timing" OR "episode" OR "occasion" OR "synchronicity" OR "event*" OR "moment" OR "periodicity" OR "tempo" OR "synchronization" OR "synchronsation" OR "frequency" OR "famil*"))<br>AND<br>("Afghanistan" OR "Bangladesh" OR "Bhutan" OR "India" OR "Maldives" OR "Nepal" OR "Sri Lanka" OR "Pakistan") NOT ("minimum meal frequency" OR "food frequency questionnaire") |

**Table S2:** Geographical, population, and methodological characteristics of included studies.

| Study reference | Country | Study population | Data collection methods | Food choice behavior(s) assessed | Domains of food choice driver(s) assessed |
| --- | --- | --- | --- | --- | --- |
| Aggarwal et al. 2006 | India | Adolescents (n=500 females and 500 males) | Interviewer-administered questionnaire | Household consumption practices | Intrapersonal, socio-cultural, personal food environment |
| Ali et al. 2021 | India | Adults (n=66 females and 122 males) | Interviewer-administered questionnaire | Food acquisition | Intrapersonal, personal food environment, material assets and resources |
| Arora et al. 2021 | India | Adults (n=102 females and 204 males) | Interviewer-administered questionnaire | Food acquisition | Intrapersonal, socio-cultural, personal food environment |
| Ashraf 2019 | Bangladesh | Adolescents and adults (n=37 females and 63 males) | Interviewer-administered questionnaire | Food acquisition | Intrapersonal, material assets and resources |
| Bailey et al. 2018 | India | Adults (n=38 females) | Semi-structured interviews, pile sorting | Food preparation, Household consumption practices | Intrapersonal, socio-cultural, personal food environment, person-state |
| Bhol et al. 2021 | India | Adults (n=196 females and 60 males) | Self-administered questionnaire | Household consumption practices | Socio-cultural, person-state |
| Blum et al. 2019 | Bangladesh | Adolescents (n=24) | Semi-structured interviews | Household consumption practices | Socio-cultural, personal food environment |
| Budhathoki, M., & Pandey, S. 2021 | Nepal | Adults (n=304 females and 224 males) | Interviewer-administered questionnaire | Food acquisition | Intrapersonal, socio-cultural, personal food environment |
| Bukhari et al. 2022 | Pakistan | Adults (n=46 females and 44 males) | Semi-structured interviews | Food acquisition | Socio-cultural, personal food environment, person-state |
| Chakrabarti, S. 2010 | India | Adults (n=33 experts) | Interviewer-administered questionnaire | Food acquisition | Intrapersonal, socio-cultural, personal food environment |
| Chopra et al. 2021 | India | Adolescents (n=20 females and 16 males) and adult caregivers (n=23 females) | Focus group discussions (FGDs) | Food acquisition, Food preparation, Household consumption practices | Intrapersonal, socio-cultural, personal food environment |
| Dahal et al. 2022 | Nepal | Adolescent and adult students (n=194 females and 191 males) | Self-administered questionnaire | Household consumption practices | Intrapersonal, socio-cultural, personal food environment |
| Das et al. 2021 | India | Adults (n=68 females and 82 males) | Self-administered questionnaire, semi-structured interviews | Food acquisition, Food preparation | Intrapersonal, person-state |
| David et al. 2020 | India | Adults (n=112 females and 128 males) | Interviewer-administered questionnaire | Food acquisition | Intrapersonal |
| Ganpule et al. 2023 | India | Adults (n=4949 females and 3813 males) | Interviewer-administered questionnaire | Household consumption practices | Intrapersonal, material assets and resources |
| Ghatak et al. 2023 | India | Adults (n=1125 households each with at least one female and one male aged up to 18 years) | Interviewer-administered questionnaire, photovoice | Household consumption practices | Socio-cultural, material assets and resources |
| Kabir et al. 2018 | Bangladesh | Adolescents and adults (Interviews: n=9 females and 19 males, FGDs: n=10 females and 16 males) | Semi-structured interviews, focus group discussions | Food preparation, Household consumption practices | Intrapersonal, socio-cultural, material assets and resources |
| Kaur et al. 2020 | India | Adults (n=30 females and 23 males) | Focus group discussions | Household consumption practices | Intrapersonal, personal food environment |
| Khan et al. 2021 | India | Adult consumers (n=54 females and 103 males) | Interviewer-administered questionnaire | Household consumption practices | Intrapersonal |

| **Study reference** | **Country** | **Study population** | **Data collection methods** | **Food choice behavior(s) assessed** | **Domains of food choice driver(s) assessed** |
| --- | --- | --- | --- | --- | --- |
| Khanna et al. 2022 | India | Adult consumers (n=240 females and 205 males) | Self-administered questionnaire | Household consumption practices | Intrapersonal, socio-cultural, personal food environment, person-state |
| Khongrangjem et al. 2018 | India | Adolescent and adult Pre-University College students (n=77 females and 83 males) | Self-administered questionnaire | Household consumption practices | Intrapersonal |
| Kiran et al. 2018 | India | Adolescents and adults (n=66 females and 194 males) | Interviewer-administered questionnaire | Food acquisition | Intrapersonal, personal food environment |
| Kumar et al. 2017 | India | School-going adolescents (n=885 females and 767 males) | Self-administered questionnaire | Household consumption practices | Intrapersonal |
| Malushte et al. 2022 | India | Adolescents (n=96 females and 104 males) | Interviewer-administered questionnaire | Food acquisition | Intrapersonal, socio-cultural, personal food environment |
| Matharu et al. 2022 | India | Adult higher education students (n=186 females and 215 males) | Self-administered questionnaire | Food acquisition | Intrapersonal, socio-cultural, personal food environment |
| Matharu et al. 2023 | India | Adult higher education students (n=186 females and 215 males) | Self-administered questionnaire | Food acquisition | Intrapersonal, personal food environment |
| Menon et al. 2022 | India | Adult female food gatekeepers/ those responsible for feeding their family members (n=34) | Semi-structured interviews | Food acquisition, Food preparation, Household consumption practices | Socio-cultural |
| Mohanty et al. 2017 | India | Adult medical students (n=126 females and 152 males) | Interviewer-administered questionnaire | Household consumption practices | Intrapersonal, personal food environment |
| Moitra, P. & Madan, J. 2022 | India | Adolescents (n=343 females and 369 males) | Self-administered questionnaire | Food acquisition, Household consumption practices | Intrapersonal, personal food environment |
| Mor, K., & Sethia, S. 2018 | India | Adults (n=1421) | Interviewer-administered questionnaire | Household consumption practices | Intrapersonal, personal food environment |
| Neogy, S. 2010 | India | Adults (families in the community) | Focus group discussions, pile sorting | Household consumption practices | Intrapersonal, socio-cultural |
| Patel et al. 2010 | India | Adolescents (n=101 females and 50 males) | Interviewer-administered questionnaire | Household consumption practices | Intrapersonal, socio-cultural, personal food environment |
| Pradhan et al. 2013 | India | Adults (20 households and 4 key informants) | Interviewer-administered questionnaire, semi-structured interviews | Food acquisition, Household consumption practices | Intrapersonal |
| Prasad, R., & Umesh, S. 2016 | India | Not specified (n=171 respondents) | Self-administered questionnaire | Food acquisition | Intrapersonal, personal food environment |
| Qazi et al. 2022 | Pakistan | Adult students (n=36 females and 84 males) | Semi-structured interviews | Food acquisition | Socio-cultural, personal food environment, person-state |
| Rahman et al. 2021 | India | Not specified (n=416 respondents) | Self-administered questionnaire | Food acquisition | Intrapersonal, personal food environment |
| Renzella et al. 2020 | Sri Lanka | Adults (n=62 females and 32 males) | Semi-structured interviews | Food preparation | Intrapersonal, socio-cultural, material assets and resources |
| Roy et al. 2021 | India | Younger children, adolescents, and adults (n=2355 females and 2254 males) | Semi-structured interviews | Household consumption practices | Intrapersonal, personal food environment, material assets and resources, person-state |
| Samanta et al. 2022 | India | Adults (n=741 females and 318 males) | Self-administered questionnaire | Food acquisition, Food preparation | Intrapersonal, socio-cultural, person-state |

| **Study reference** | **Country** | **Study population** | **Data collection methods** | **Food choice behavior(s) assessed** | **Domains of food choice driver(s) assessed** |
| --- | --- | --- | --- | --- | --- |
| Semar, R., & Bakshi, N. 2022 | India | School-going younger children (n=73 females and 27 males) | Self-administered questionnaire | Household consumption practices | Intrapersonal, personal food environment, person-state |
| Shahzad et al. 2022 | Pakistan | Adults (n=430 females and 637 males) | Self-administered questionnaire, photovoice | Food acquisition | Socio-cultural |
| Turner et al. 2022 | India | Adults (n=16 females and 20 males) | Semi-structured interviews, photovoice | Food acquisition, Food preparation, Household consumption practices | Personal food environment |
| Ullah et al. 2023 | Pakistan | Adults (46 females and 44 males) | Semi-structured interviews | Food acquisition | Intrapersonal, socio-cultural, personal food environment |
| Verma et al. 2023 | India | Adolescents and adults (n=351 female, 371 male children and their parents) | Interviewer-administered questionnaire | Household consumption practices | Intrapersonal, socio-cultural |
| Vinish et al. 2021 | India | Adults (n=385) | Self-administered questionnaire, Interviewer-administered questionnaire | Food acquisition | Intrapersonal, personal food environment, |
| Ynion et al. 2021 | India | Adults (n=501) | Interviewer-administered questionnaire, focus group discussions | Food acquisition, Household consumption practices | Intrapersonal, personal food environment |

